# Aspirin and healthy longevity within racial and ethnic minoritized older adults in the United States

**DOI:** 10.64898/2026.08.24.26361036

**Authors:** George Tzimas, Joseph C. Vanghelof, Aseel Mohammed, Daniela S. Raicu, Lianlian Du, Michael E. Ernst, Erica T. Warner, Andrew T. Chan, Joanne Ryan, Sara E. Espinoza, Anne Murray, Kerry Sheets, Roselyne B. Tchoua, Raj C. Shah

## Abstract

**Importance:** The ASPREE randomized trial found no overall benefit of low-dose aspirin for disability-free survival among older adults. However, individual estimates in pre-specified subgroups indicated potential benefit among racial and ethnic minoritized participants in the United States (US).

**Objective:** To evaluate whether the effect of low-dose aspirin vs placebo on disability-free survival differed across US Black and Hispanic ASPREE participants using individualized treatment-effect estimation.

**Design, Setting, and Participants:** Post hoc clinical trial analysis of ASPREE, a randomized, double-blind, placebo-controlled clinical trial of daily low-dose aspirin vs placebo. This analysis included US ASPREE participants who self-identified as non-Hispanic Black or Hispanic, were aged 65 years or older, and had complete baseline predictor and outcome data.

**Interventions:** Randomization to daily 100-mg aspirin or placebo.

**Main Outcomes and Measures:** The primary outcome was loss of disability-free survival, defined as death, persistent physical disability, or dementia. Individualized treatment effects were estimated post hoc using a Random Survival Forest X-learner. Heterogeneity was evaluated on the relative scale with Cox proportional hazards models and on the absolute scale with 5-year risk differences.

**Results:** Among 2411 US ASPREE participants, 1270 were included in the Black and Hispanic analytic cohort (897 non-Hispanic Black and 373 Hispanic participants; mean age, 71.8 years). Aspirin was associated with lower risk of disability-free survival loss compared with placebo (hazard ratio [HR], 0.65; 95% CI, 0.45-0.93). In model-derived tertiles, aspirin was associated with lower risk in the greatest predicted-benefit group (HR, 0.36; 95% CI, 0.19-0.71; 5-year absolute risk difference [ARD], −11.1 percentage points; 95% CI, −22.0 to −0.1) but not in the lowest predicted-benefit group (HR, 1.26; 95% CI, 0.70-2.27; ARD, +3.9 percentage points; 95% CI, −5.9 to 13.6).

**Conclusions and Relevance:** In these analyses of US Black and Hispanic ASPREE participants, aspirin effects on disability-free survival appear to be heterogeneous, with benefit concentrated in a subset of participants. Because these findings are from post-hoc models, they should be externally validated before being incorporated into clinical decision-making.

**Trial Registration:** ClinicalTrials.gov Identifier: <u>NCT01038583</u>; https://clinicaltrials.gov/study/NCT01038583

## Introduction

In the United States (US), both the total number and proportion of older adults continue to grow, with increasing racial and ethnic diversity among adults over age 65 (1). Racial and ethnic minoritized communities face a disproportionate burden of chronic disease, disability risk, and barriers to healthy aging (2). Improving health outcomes within these populations (i.e. minority health) requires understanding how preventive interventions work within groups rather than only comparing effects between groups (3).

Daily low-dose aspirin has been studied for its potential to extend a healthy longevity in older adults (4). The ASPREE randomized clinical trial (RCT) of adults aged 70 and older (or 65 and older for Black and Hispanic participants) reported that aspirin did not improve disability-free survival in the overall population (5). Additional analyses showed no reduction in cardiovascular events, increased major hemorrhage, and higher all-cause mortality with aspirin (6,7). ASPREE findings factored into the US Preventive Services Task Force recommendation “with moderate certainty that initiating aspirin use for the primary prevention of CVD events in adults 60 years or older has no net benefit” (8). Contemporary ACC/AHA primary prevention guidance similarly discourages routine aspirin use for primary prevention among adults older than 70 years or those at increased bleeding risk (9). Although overall group interaction by race/ethnicity in ASPREE was not significant, favorable individual point estimates were observed for certain US minoritized groups, raising the possibility of subgroup-specific treatment effects (5).

Traditional subgroup analyses have limited ability to detect meaningful patterns of heterogeneity of treatment effect (HTE), especially when treatment effects differ among individuals with varying functional, cognitive, and clinical profiles (10). Because these methods typically depend on pre-specified interactions or broadly defined risk groups, they may overlook more complex, nonlinear variations in treatment response. Modern approaches to HTE, which estimate how intervention effects differ across individuals, are therefore needed to revisit earlier findings with updated modeling frameworks (10,11). To address these limitations, this post hoc analysis used an individualized treatment-effect framework to estimate participant-specific conditional average treatment effects (CATEs) for aspirin vs placebo among US Black and Hispanic ASPREE participants.

## Methods

### Study Population

The Institutional Review Board of The University of Iowa provided ethical approval for this work which was part of the ASPREE-XT study including analyses of data from the ASPREE randomized clinical trial. We analyzed US participant data from ASPREE, a randomized, double-blind, parallel-group, placebo-controlled trial evaluating daily 100 mg aspirin versus placebo among community-dwelling older adults free from cardiovascular disease, dementia, or persistent physical disability at baseline (5). US racial and ethnic minoritized participants were defined as individuals self-identifying as non-Hispanic Black (hereafter referred to as Black) or Hispanic. The final analytic cohort included 1,270 minoritized participants.

### Data Preparation

The primary endpoint was time to loss of disability-free survival, defined as the first occurrence of dementia, persistent physical disability, or death. Baseline predictors were a collection of demographic, clinical, laboratory, medication, functional, cognitive, and depressive symptom measures collected before randomization. Definitions and preprocessing are provided in the eMethods.

### Individualized Treatment Effect Estimation

The analytic workflow for estimating individualized treatment effects is shown in eFigure 1. For each participant, the conditional average treatment effect (CATE) was defined as the estimated 5-year risk of disability-free survival loss if assigned to aspirin minus the estimated risk if assigned to placebo. Negative values indicated predicted benefit.

**Figure 1.**
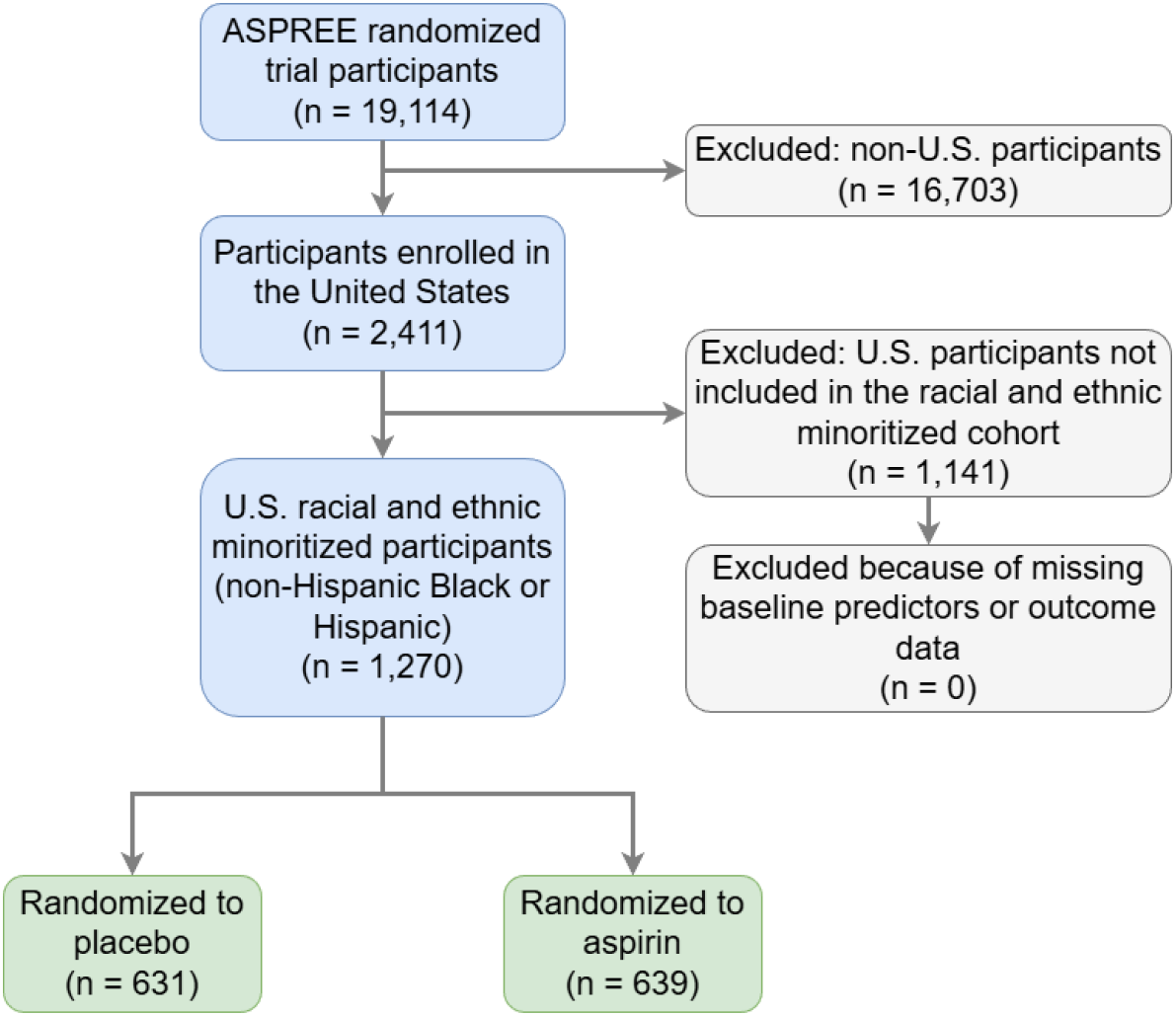
Study population flow diagram.

**Figure 2.**
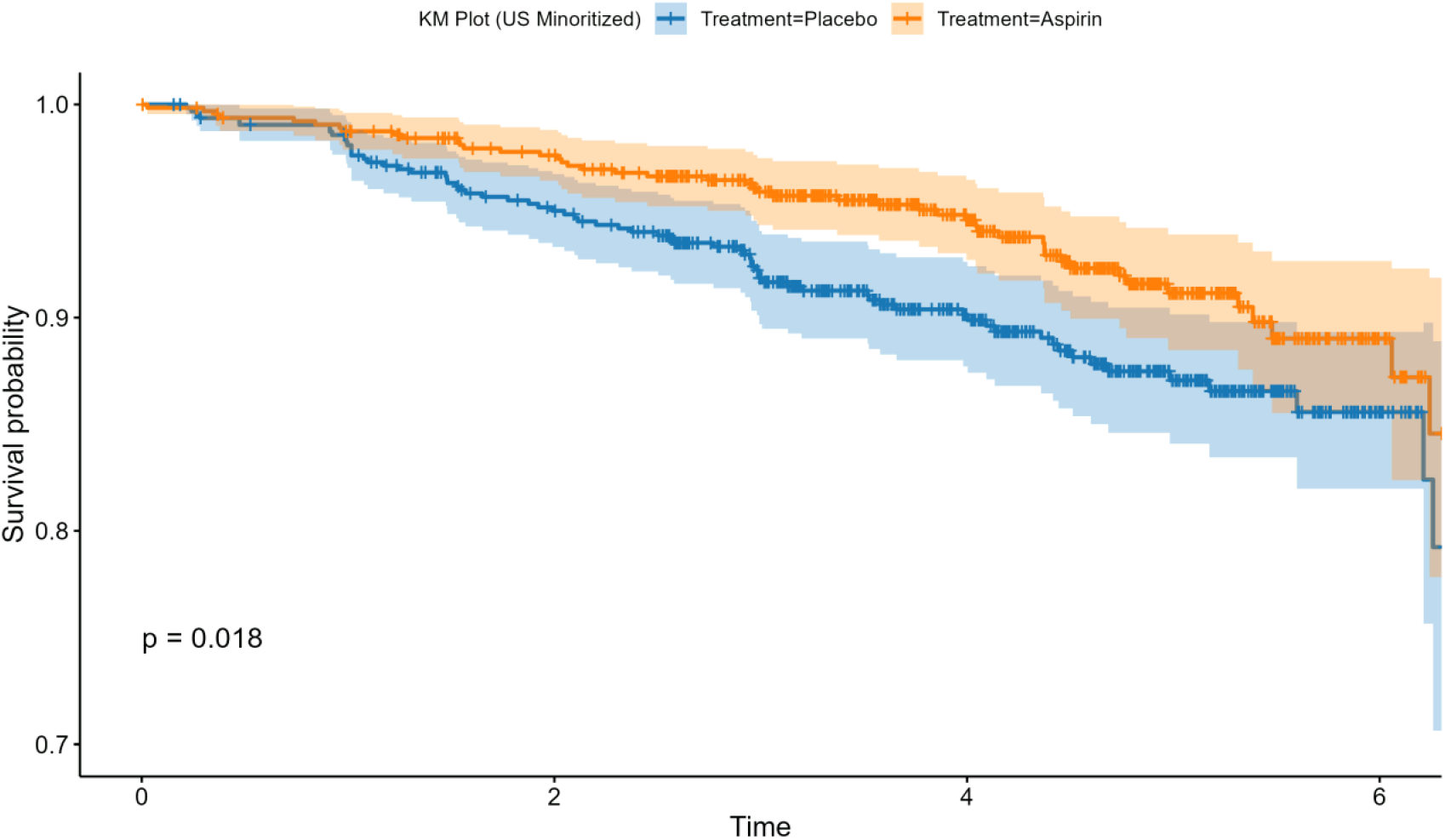
Kaplan-Meier survival curves for the US Black and Hispanic cohort comparing aspirin versus placebo. The p-value shown corresponds to the log-rank test.

CATEs were estimated using X-learner, a meta-learning framework for estimating heterogeneous treatment effects, using arm-specific Random Survival Forest outcome models and random forest treatment-effect models (12–14). Five-fold cross-fitting was used to generate predictions, with separate fold assignments for the outcome-model and treatment-effect-model stages to reduce overfitting and data leakage across the modeling steps (15). Inverse probability of censoring weights from cross-fitted ranger censoring models were used in the pseudo-outcome stage to reduce bias from incomplete 5-year outcome ascertainment due to censoring (16). Predictor definitions, sensitivity analyses, and model parameters are provided in the eMethods and eTables 1-2.

### Statistical Analyses

Heterogeneity of treatment effect was evaluated by testing treatment interactions with standardized continuous CATE and with model-derived CATE tertiles (T1, greatest predicted benefit; T3, lowest predicted benefit or possible harm). Relative-scale analyses used Cox proportional hazards models and likelihood ratio tests; absolute-scale analyses used linear models for the 5-year event indicator and analysis of variance. Kaplan-Meier estimates were descriptive. Baseline characteristics across CATE groups were summarized descriptively. Sensitivity analyses repeated the X-learner using random forests for the arm-specific outcome stage, and computational stability was assessed by refitting the primary model with 10 additional random seeds (eTable 3).

## Results

### Participant Characteristics

Figure 1 shows the flow of participants into these analyses. Among the 2,411 US participants, 897 identified as Black and 373 as Hispanic. In the combined Black and Hispanic cohort (n = 1270), the mean age was 71.8 years and overweight or obesity was common (82.3%). Key baseline characteristics were similarly distributed by aspirin vs placebo group (Table 1).

**Table 1.** Baseline characteristics of US racial and ethnic minoritized participants by treatment group.

| Variable | Aspirin<sup>1</sup><br>(n=639) | Placebo<sup>1</sup><br>(n=631) | p<sup>2</sup> |
| --- | --- | --- | --- |
| Age (years) | 71.87 (5.33) | 71.68 (5.48) | 0.53 |
| Race/Ethnicity |  |  | 0.73 |
| Black | 448 (70.1%) | 449 (71.2%) |  |
| Hispanic | 191 (29.9%) | 182 (28.8%) |  |
| Education level |  |  | 0.62 |
| <9 yrs | 50 (7.8%) | 62 (9.8%) |  |
| 9-11 yrs | 54 (8.5%) | 54 (8.6%) |  |
| 12 yrs | 159 (24.9%) | 147 (23.3%) |  |

| **Variable** | **Aspirin<sup>1</sup>**<br>**(n=639)** | **Placebo<sup>1</sup>**<br>**(n=631)** | **p<sup>2</sup>** |
| --- | --- | --- | --- |
| 13-15 yrs | 197 (30.8%) | 176 (27.9%) |  |
| 16 yrs | 85 (13.3%) | 87 (13.8%) |  |
| 17-21 yrs | 94 (14.7%) | 105 (16.6%) |  |
| Living situation |  |  | 0.26 |
| At home alone | 248 (38.8%) | 267 (42.3%) |  |
| At home with family,<br>friends or spouse | 387 (60.6%) | 357 (56.6%) |  |
| In a residential home<br>(supervised care) | 4 (0.6%) | 7 (1.1%) |  |
| In a nursing home<br>alone | 0 (0.0%) | 0 (0.0%) |  |
| Smoking status |  |  | 0.40 |
| Current | 66 (10.3%) | 76 (12.0%) |  |
| Former | 244 (38.2%) | 221 (35.0%) |  |
| Never | 329 (51.5%) | 334 (52.9%) |  |
| Alcohol use |  |  | 0.95 |
| Current | 299 (46.8%) | 299 (47.4%) |  |
| Former | 120 (18.8%) | 114 (18.1%) |  |
| Never | 220 (34.4%) | 218 (34.5%) |  |
| Body mass index<br>(kg/m<sup>2</sup>) |  |  | 0.80 |
| Underweight | 4 (0.6%) | 2 (0.3%) |  |
| Normal | 106 (16.6%) | 113 (17.9%) |  |
| Overweight | 243 (38.0%) | 237 (37.6%) |  |
| Obese | 286 (44.8%) | 279 (44.2%) |  |
| Abdominal circumference (cm) | 100.06 (14.37) | 100.09 (13.66) | 0.97 |
| Diabetes | 152 (23.8%) | 166 (26.3%) | 0.33 |
| Family history of heart attack | 110 (17.2%) | 105 (16.6%) | 0.84 |
| Systolic blood pressure (mmHg) | 136.33 (17.10) | 136.16 (17.12) | 0.86 |
| Diastolic blood pressure (mmHg) | 79.41 (9.70) | 79.32 (9.99) | 0.88 |
| Average dominant grip strength | 26.99 (12.13) | 26.86 (12.41) | 0.85 |
| Average gait speed (s/5m) | 4.19 (1.90) | 4.24 (2.00) | 0.69 |
| 3MS overall score | 91.69 (5.19) | 91.97 (5.26) | 0.33 |
| CES-D overall score | 3.59 (3.55) | 3.85 (4.02) | 0.22 |
| HDL cholesterol (mmol/L) | 1.51 (0.42) | 1.55 (0.45) | 0.11 |
| LDL cholesterol (mmol/L) | 2.84 (0.86) | 2.87 (0.92) | 0.50 |
| eGFR (CKD) | 78.07 (17.17) | 78.48 (17.90) | 0.67 |
| Hemoglobin (g/dL) | 13.53 (1.35) | 13.49 (1.36) | 0.59 |
| Use of antihypertensive medications | 39 (6.1%) | 29 (4.6%) | 0.28 |
| Use of lipid-lowering agents | 350 (54.8%) | 347 (55.0%) | 0.98 |
| <sup>1</sup> Mean (SD); n (%) |  |  |  |
| <sup>2</sup> Welch Two Sample t-test; Pearson's Chi-squared test |  |  |  |

### Aspirin and Disability-Free Survival in Racial and Ethnic Minoritized Participants

Aspirin was associated with a lower rate of disability-free survival loss. The unadjusted Kaplan-Meier curves showed separation between the aspirin and placebo arms (log-rank P = .018; Figure 2), and the corresponding Cox proportional hazards model estimated an HR of 0.65 (95% CI, 0.45-0.93). In contrast, no evidence of benefit was observed among US non-Hispanic White participants, with overlapping Kaplan-Meier curves (log-rank P = .46) and a Cox model HR of 1.04 (95% CI, 0.94-1.14) (eFigure 2).

**Figure 2.**
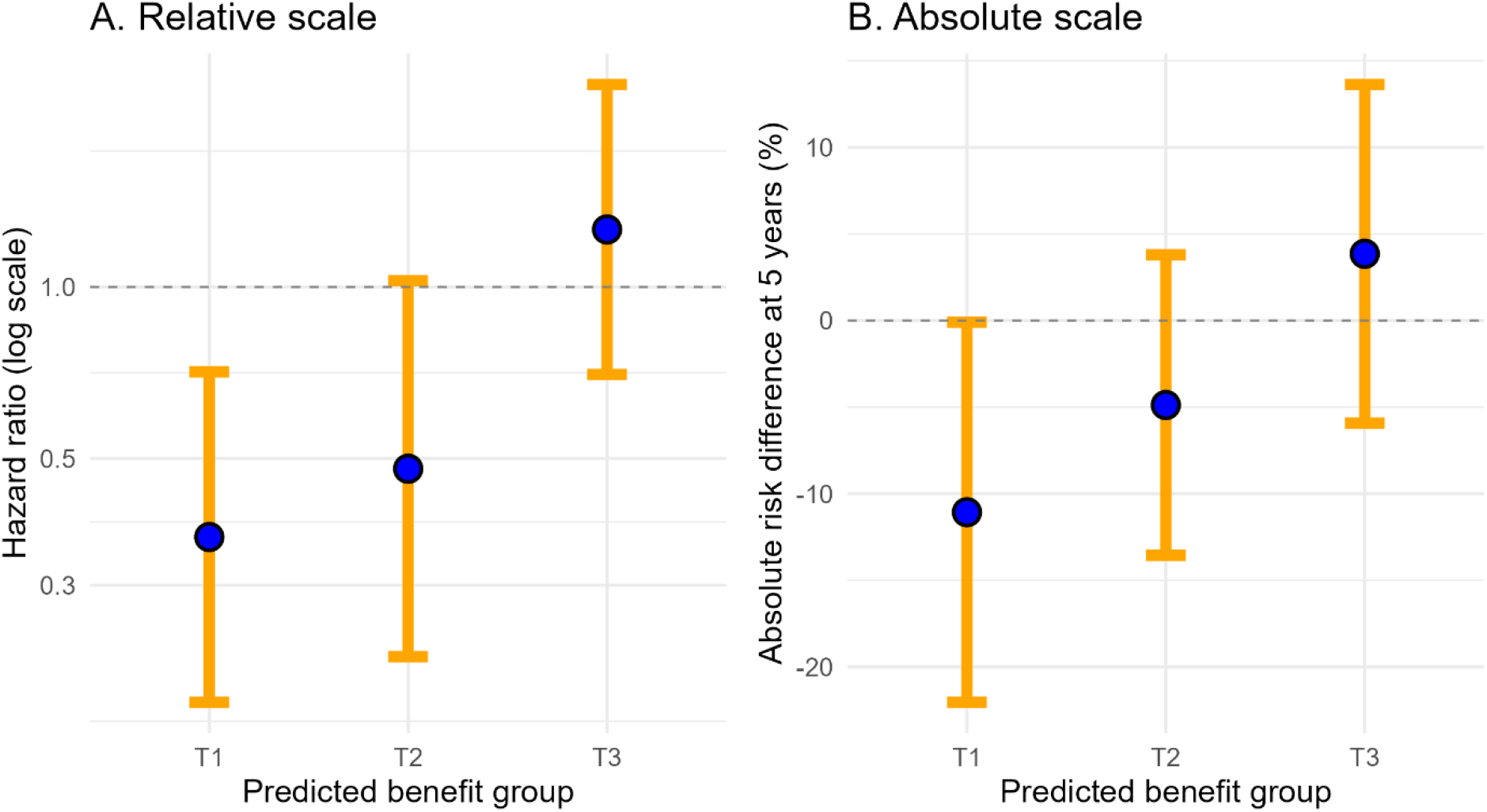
Relative and absolute treatment effects of aspirin by predicted benefit group. Panel A shows hazard ratios for aspirin versus placebo for loss of disability-free survival across model-derived predicted benefit tertiles. Panel B shows 5-year absolute risk differences for aspirin minus placebo across predicted benefit tertiles. T1 represents the greatest predicted benefit from aspirin, and T3 represents the lowest predicted benefit or possible harm. Hazard ratios less than 1.0 and absolute risk differences less than 0 favor aspirin.

### Individualized Treatment Effects and Heterogeneity of Response

Using the RSF-based X-learner, estimated CATE values varied across participants and were generally centered below zero, consistent with lower predicted 5-year risk of disability-free survival loss under aspirin on average (eFigure 3). Discrimination at the 5-year horizon was modest (eTable 4). As shown in eTable 5, the continuous CATE-by-treatment interaction was statistically significant on the relative scale (interaction HR per 1-SD higher CATE, 1.82; 95% CI, 1.28-2.60; likelihood ratio P = .001) and on the absolute scale (ANOVA interaction P = .011).

Kaplan-Meier curves stratified by model-derived CATE tertile and treatment assignment are shown in eFigure 4. Relative and absolute treatment-effect estimates are summarized in Figure 3. The relative-scale tertile-by-treatment interaction was significant (likelihood ratio P = .011). In T1, aspirin was associated with a lower hazard of the primary outcome (HR, 0.36; 95% CI, 0.19-0.71; P = .003). In T2, the estimated association was smaller and did not reach statistical significance (HR, 0.48; 95% CI, 0.22-1.03; P = .058), and there was no evidence of benefit in T3 (HR, 1.26; 95% CI, 0.70-2.27; P = .44). The absolute-scale tertile-by-treatment interaction was also significant (ANOVA P = .036). Aspirin was associated with an 11.1-percentage-point reduction in 5-year risk in T1 (ARD, −11.1; 95% CI, −22.0 to − 0.1; P = .048), whereas estimates in T2 (ARD, −4.9; 95% CI, −13.6 to 3.8; P = .269) and T3 (ARD, +3.9; 95% CI, −5.9 to 13.6; P = .438) were compatible with no benefit.

The RF-based sensitivity analysis showed a similar pattern, with benefit concentrated in T1 and no clear benefit in T3 (eTable 6). Continuous interactions remained significant on the relative (P = .002) and absolute (P = .006) scales, as did tertile-by-treatment interactions (relative P = .025; absolute P = .034). In T1, RF-based estimates were HR, 0.33 (95% CI, 0.18-0.62) and ARD, −13.7 percentage points (95% CI, −24.5 to −2.9); in T3, they were HR, 1.04 (95% CI, 0.55-1.97) and ARD, +1.6 percentage points (95% CI, −8.1 to 11.4).

### Baseline Characteristics Across Model-Derived CATE Groups

Baseline characteristics across the three model-derived CATE groups are shown in Table 2. Differences in demographic, clinical, and functional characteristics across groups suggested that the model-derived strata captured important variation in participant profiles.

**Table 2.** Baseline characteristics of participants stratified by predicted CATE group (T1–T3).

| Variable | T1<sup>1</sup> | T2<sup>1</sup> | T3<sup>1</sup> | p<sup>2</sup> |
| --- | --- | --- | --- | --- |
| Age (years) | 70.88 (5.17) | 71.60 (4.95) | 72.84 (5.88) | <0.001 |
| Race/Ethnicity |  |  |  | 0.52 |
| Black | 308 (72.6%) | 293 (69.3%) | 296 (70.0%) |  |
| Hispanic | 116 (27.4%) | 130 (30.7%) | 127 (30.0%) |  |
| Education level |  |  |  | 0.007 |
| <9 yrs | 34 (8.0%) | 34 (8.0%) | 44 (10.4%) |  |
| 9-11 yrs | 39 (9.2%) | 35 (8.3%) | 34 (8.0%) |  |
| 12 yrs | 127 (30.0%) | 105 (24.8%) | 74 (17.5%) |  |
| 13-15 yrs | 112 (26.4%) | 132 (31.2%) | 129 (30.5%) |  |
| 16 yrs | 49 (11.6%) | 61 (14.4%) | 62 (14.7%) |  |
| 17-21 yrs | 63 (14.9%) | 56 (13.2%) | 80 (18.9%) |  |

| **Variable** | **T1<sup>1</sup>** | **T2<sup>1</sup>** | **T3<sup>1</sup>** | **p<sup>2</sup>** |
| --- | --- | --- | --- | --- |
| Living situation |  |  |  | 0.006 |
| At home alone | 201 (47.4%) | 161 (38.1%) | 153 (36.2%) |  |
| At home with family, friends or spouse | 221 (52.1%) | 259 (61.2%) | 264 (62.4%) |  |
| In a residential home (supervised care) | 2 (0.5%) | 3 (0.7%) | 6 (1.4%) |  |
| Smoking status |  |  |  | <0.001 |
| Current | 96 (22.6%) | 32 (7.6%) | 14 (3.3%) |  |
| Former | 131 (30.9%) | 163 (38.5%) | 171 (40.4%) |  |
| Never | 197 (46.5%) | 228 (53.9%) | 238 (56.3%) |  |
| Alcohol use |  |  |  | 0.32 |
| Current | 205 (48.3%) | 193 (45.6%) | 200 (47.3%) |  |
| Former | 83 (19.6%) | 85 (20.1%) | 66 (15.6%) |  |
| Never | 136 (32.1%) | 145 (34.3%) | 157 (37.1%) |  |
| Body mass index (kg/m<sup>2</sup>) |  |  |  | 0.009 |
| Underweight | 2 (0.5%) | 1 (0.2%) | 3 (0.7%) |  |
| Normal | 64 (15.1%) | 65 (15.4%) | 90 (21.3%) |  |
| Overweight | 152 (35.8%) | 154 (36.4%) | 174 (41.1%) |  |
| Obese | 206 (48.6%) | 203 (48.0%) | 156 (36.9%) |  |
| Abdominal circumference (cm) | 101.78 (14.21) | 100.62 (12.31) | 97.82 (15.12) | <0.001 |
| Diabetes | 131 (30.9%) | 103 (24.3%) | 84 (19.9%) | <0.001 |
| Family history of heart attack | 70 (16.5%) | 62 (14.7%) | 83 (19.6%) | 0.15 |
| Systolic blood pressure (mmHg) | 138.13 (16.76) | 134.63 (16.29) | 135.98 (18.07) | 0.008 |
| Diastolic blood pressure (mmHg) | 82.04 (10.62) | 78.60 (8.79) | 77.45 (9.45) | <0.001 |
| Average dominant grip strength | 28.05 (12.53) | 27.57 (12.36) | 25.15 (11.73) | <0.001 |
| Average gait speed (s/5m) | 4.36 (2.27) | 4.08 (1.74) | 4.21 (1.80) | 0.13 |
| 3MS overall score | 91.59 (5.30) | 92.15 (4.99) | 91.75 (5.38) | 0.26 |
| CES-D overall score | 3.97 (4.27) | 3.59 (3.38) | 3.58 (3.66) | 0.28 |
| HDL cholesterol (mmol/L) | 1.40 (0.36) | 1.50 (0.36) | 1.69 (0.52) | <0.001 |
| LDL cholesterol (mmol/L) | 2.71 (0.93) | 2.89 (0.82) | 2.97 (0.90) | <0.001 |
| eGFR (CKD) | 78.10 (20.22) | 77.98 (16.06) | 78.74 (16.00) | 0.77 |
| Hemoglobin (g/dL) | 13.98 (1.55) | 13.47 (1.24) | 13.10 (1.09) | <0.001 |
| Use of antihypertensive medications | 35 (8.3%) | 20 (4.7%) | 13 (3.1%) | 0.003 |
| Use of lipid-lowering agents | 227 (53.5%) | 224 (53.0%) | 246 (58.2%) | 0.25 |

<sup>1</sup>Mean (SD); n (%)
<sup>2</sup>One-way analysis of means (not assuming equal variances); Pearson's Chi-squared test

Compared with T3, participants in T1 were younger and more often lived alone, currently smoked, had diabetes or obesity, and used antihypertensive medication. They also had higher abdominal circumference, systolic and diastolic blood pressure, grip strength, and hemoglobin, with lower HDL and LDL cholesterol.

## Discussion

In this post hoc secondary analysis of US Black and Hispanic older adults from the ASPREE randomized clinical trial, aspirin compared with placebo was associated with a lower hazard of incident persistent disability, dementia, or death. Individualized treatment-effect modeling suggested that this association was not uniform across participants: heterogeneity was observed on both relative and absolute scales, and benefit was concentrated in the model-derived group with the greatest predicted benefit.

The greatest predicted-benefit group had lower relative hazard and lower 5-year absolute risk with aspirin, whereas the lowest predicted-benefit group had no clear evidence of benefit. These findings suggest that average treatment effects may mask clinically meaningful differences in aspirin response, particularly among populations that have historically been underrepresented in prevention trials.

The original ASPREE trial found no overall benefit of aspirin in the full study population, while subgroup estimates suggested possible differences among US minoritized participants. This analysis extends that observation by using a multivariable X-learner framework to explore nonlinear patterns of treatment-effect heterogeneity rather than one-variable-at-a-time subgroup contrasts.

The factors underlying the observed variation in predicted aspirin benefit remain uncertain. Differences across CATE groups were not explained simply by race or ethnicity within the analytic cohort. Instead, the greatest predicted-benefit group had a profile characterized by higher cardiometabolic risk, including more current smoking, diabetes, obesity, higher blood pressure, and greater antihypertensive medication use, together with higher grip strength and hemoglobin and lower HDL and LDL cholesterol. These comparisons are descriptive and hypothesis-generating, not evidence that any individual characteristic modifies aspirin’s effect.

Strengths include randomized treatment assignment, adjudicated trial outcomes, and individualized treatment-effect estimation, which can capture more complex HTE patterns than conventional subgroup analyses. The primary pattern was also directionally consistent in a random forest sensitivity analysis and in seed-based stability checks.

Several limitations should be considered. The analysis was post hoc and not prespecified in the original ASPREE protocol. The sample size was modest for treatment-effect heterogeneity modeling, and the model-derived CATE strata and cut points require external validation. Individualized treatment effects were predictions based on measured baseline covariates instead of being directly observed individual causal effects. Finally, the lack of an independent validation cohort limits generalizability and precludes clinical use of the CATE groups at this stage.

## Conclusion

Among US Black and Hispanic ASPREE participants, these analyses suggest heterogeneity in the estimated effect of aspirin on disability-free survival, with benefit concentrated in a subset of participants and little or no apparent benefit in others. These post-hoc modelling results require validation in independent data before being used to guide preventive aspirin decisions.

## Data Sharing Statement

Deidentified participant data and a data dictionary may be available to qualified researchers after approval of a proposal and completion of a data access agreement through ASPREE investigators; details are available at https://aspree.org/usa/for-researchers/ and https://aspree.org/aus/for-researchers/.

## Author Contributions

Mr Tzimas and Dr Shah had full access to all the data in the study and take responsibility for the integrity of the data and the accuracy of the data analysis.

Concept and design: Tzimas, Vanghelof, Mohammed, Tchoua, Shah. Acquisition, analysis, or interpretation of data: All authors.

Drafting of the manuscript: Tzimas, Tchoua, Shah.

Critical review of the manuscript for important intellectual content: All authors. Statistical analysis: Tzimas, Vanghelof.

Obtained funding: Chan, Ryan, Murray.

Administrative, technical, or material support: Raicu, Ernst, Warner, Chan, Ryan, Espinoza, Murray, Sheets, Shah.

Supervision: Tchoua, Shah.

## Acknowledgements

The authors recognize the significant contributions made by the research participants, staff, and investigators for the ASPirin in Reducing Events in the Elderly clinical trial.

## References

1. Administration for Community Living. 2023 Profile of Older Americans [Internet]. Washington, DC: U.S. Department of Health and Human Services; 2024. Available from: https://acl.gov/sites/default/files/Profile%20of%20OA/ACL_ProfileOlderAmericans202 3_508.pdf

2. National Academies of Sciences E, Division H and M, Practice B on PH and PH, States C on CBS to PHE in the U, Baciu A, Negussie Y, et al. The State of Health Disparities in the United States. In: Communities in Action: Pathways to Health Equity [Internet]. National Academies Press (US); 2017 [cited 2025 Dec 11]. Available from: https://www.ncbi.nlm.nih.gov/books/NBK425844/

3. Whitfield KE, Allaire JC, Belue R, Edwards CL. Are comparisons the answer to understanding behavioral aspects of aging in racial and ethnic groups? J Gerontol B Psychol Sci Soc Sci. 2008;63(5):P301–8.

4. ASPREE Investigator Group. Study design of ASPirin in Reducing Events in the Elderly (ASPREE): a randomized, controlled trial. Contemp Clin Trials. 2013;36(2):555–64.

5. McNeil JJ, Woods RL, Nelson MR, Reid CM, Kirpach B, Wolfe R, et al. Effect of aspirin on disability-free survival in the healthy elderly. N Engl J Med. 2018;379(16):1499–508.

6. McNeil JJ, Wolfe R, Woods RL, Tonkin AM, Donnan GA, Nelson MR, et al. Effect of aspirin on cardiovascular events and bleeding in the healthy elderly. N Engl J Med. 2018;379(16):1509–18.

7. McNeil JJ, Nelson MR, Woods RL, Lockery JE, Wolfe R, Reid CM, et al. Effect of aspirin on all-cause mortality in the healthy elderly. N Engl J Med. 2018;379(16):1519–28.

8. Davidson KW, Barry MJ, Mangione CM, Cabana M, Chelmow D, Coker TR, et al. Aspirin use to prevent cardiovascular disease: US Preventive Services Task Force recommendation statement. Jama. 2022;327(16):1577–84.

9. Coke LA, Himmelfarb CD. Guideline on the primary prevention of cardiovascular disease: let’s get it into practice! J Cardiovasc Nurs. 2019;34(4):285–8.

10. Kent DM, Steyerberg E, Van Klaveren D. Personalized evidence based medicine: predictive approaches to heterogeneous treatment effects. Bmj. 2018;363.

11. Kent DM, Paulus JK, van Klaveren D, D’Agostino R, Goodman S, Hayward R, et al. The Predictive Approaches to Treatment effect Heterogeneity (PATH) Statement. Ann Intern Med. 2020 Jan 7;172(1):35–45. doi:10.7326/M18-3667

12. Breiman L. Random forests. Mach Learn. 2001;45:5–32.

13. Ishwaran H, Kogalur UB, Blackstone EH, Lauer MS. Random survival forests. ArXiv E-Prints. 2008;arXiv-0811.

14. Künzel SR, Sekhon JS, Bickel PJ, Yu B. Metalearners for estimating heterogeneous treatment effects using machine learning. Proc Natl Acad Sci. 2019;116(10):4156–65.

15. Willems S, Schat A, Van Noorden M, Fiocco M. Correcting for dependent censoring in routine outcome monitoring data by applying the inverse probability censoring weighted estimator. Stat Methods Med Res. 2018;27(2):323–35.

16. Robins JM, Finkelstein DM. Correcting for noncompliance and dependent censoring in an AIDS clinical trial with inverse probability of censoring weighted (IPCW) log-rank tests. Biometrics. 2000;56(3):779–88.

