## Supplemental Material for "Aspirin and healthy longevity within racial and ethnic minoritized older adults in the United States"

SUPPLEMENT

### Low-dose aspirin versus placebo and healthy longevity within racial and ethnic minoritized older adults in the United States: A Post-Hoc Secondary Analysis of the ASPREE Randomized Clinical Trial

#### TABLE OF CONTENTS


eMethods. Predictor definitions, X-learner implementation, sensitivity analyses, and stability checks 2-3

eTable 1. Baseline predictors used in the primary analysis 4
eTable 2. Model parameters for the primary and sensitivity X-learners 4
eTable 3. Seed-based stability of estimated CATEs and predicted benefit tertile assignment 5
eTable 4. Discrimination of the arm-specific outcome models at the 5-year horizon 5
eTable 5. Cox proportional hazards regression results evaluating treatment, CATE, and their interaction 6
eTable 6. Sensitivity analysis comparing Random Survival Forest and Random Forest-based X-learner results 7

eFigure 1. Analytic workflow for estimating individualized treatment effects 8
eFigure 2. Distribution of estimated CATE values 9
eFigure 3. Kaplan-Meier curves by CATE tertile and treatment assignment 10
eFigure 4. Kaplan-Meier curves for US non-Hispanic White ASPREE participants 11

#### eMethods

**eMethods 1.** Predictor definitions and preprocessing
Baseline predictors for the primary analysis were selected based on clinical relevance and prior literature. The predictor set was based on previously described variables that were associated with disability-free survival and included demographic, clinical, laboratory, medication, functional, and cognitive measures which were taken at baseline.
The final set of predictors included: age; living situation; education; race/ethnicity (Black or Hispanic); diabetes status; smoking status; alcohol use; family history of heart attack; high-density lipoprotein (HDL) cholesterol; low-density lipoprotein (LDL) cholesterol; estimated glomerular filtration rate (eGFR); hemoglobin; systolic and diastolic blood pressure; body mass index (BMI) category; abdominal circumference; dominant hand grip strength; gait speed; antihypertensive medication use; lipid-lowering medication use; cognitive function (Modified Mini-Mental State Examination [3MS]); and depressive symptoms (Center for Epidemiologic Studies Depression Scale [CES-D]).

Continuous variables were analyzed on their original scale without categorization, except for the BMI category. Categorical variables, including living situation, education, smoking status, alcohol use, BMI category, and race/ethnicity and medication variables, were encoded as factors. Unordered categorical predictors were handled using partition-based splitting within tree-based models.

All analyses were restricted to US minoritized ASPREE participants included in the primary analysis. The final analytic dataset had 1,270 participants and 22 baseline predictors. The same predictor set and preprocessing steps were applied consistently for all modeling stages, including the arm-specific outcome regression models and the treatment effect estimation models.

**eMethods 2.** X-learner implementation details
We estimated individualized treatment effects using an X-learner framework adapted for time-to-event outcomes. Final estimation was the participant-specific conditional average treatment effect (CATE), defined as the predicted difference in 5-year risk of the composite endpoint under aspirin versus placebo, conditional on baseline covariates. Under this definition, more negative CATE values indicated greater predicted benefit from aspirin.
In the first stage, we fit arm-specific outcome models in the aspirin and placebo groups to estimate the 5-year risk of disability-free survival loss. The primary models on this stage were random survival forests fit using the ranger package. For each participant, the arm-specific models generated cross-fitted 5-year risks under aspirin and placebo, denoted as $\mu 1(X)$ and $\mu 0(X)$. For participants with known status under each arm, the pseudo-outcomes compared the observed event indicator with the counterfactual risk prediction from the opposite treatment arm. IPCW scores were estimated using cross-fitted ranger censoring models that included treatment and baseline predictors. These IPCW scores estimated each participant's probability of being uncensored throughout the 5-year outcome assessment, conditional on treatment and baseline predictors. The inverse of this probability was used to reweight the observed outcomes, so that participants with complete follow-up represented both themselves and similar participants who were censored. This was done in order to reduce bias from differential loss to follow-up or

incomplete outcome assessment, under the assumption that censoring was independent of the outcome conditional on measured predictors.

In the second stage, we modeled the pseudo-outcomes separately in the aspirin and placebo groups using random forest regression with IPCW weights. We used five-fold cross-fitting so that each participant's predictions were generated by models trained without that participant's fold. The two arm-specific treatment-effect predictions were equally averaged to obtain the final CATE estimates.

**eMethods 3.** Sensitivity analysis using random forests
As a sensitivity analysis, we repeated the X-learner using random forests in place of random survival forests for the arm-specific outcome regression stage, so the models estimated the binary 5-year event indicator directly. All the baseline predictors, preprocessing steps, treatment effect modeling procedures, and heterogeneity analyses remained the same. We constructed pseudo-outcomes using the random forest–based 5-year risk estimates, and we estimated participant CATE values using the same cross-fitted X-learner framework and random forest treatment effect models described above. This sensitivity analysis assessed whether the principal HTE pattern was consistent when the outcome regression modeled the 5-year binary endpoint rather than the full time-to-event outcome.

**eMethods 4.** Computational stability
To assess computational stability, the primary Random Survival Forest-based X-learner was refit with 10 additional random seeds while preserving the analytic cohort, baseline predictors, cross-fitting structure, and model specification. Agreement with the reference model was summarized with Spearman CATE correlations, exact tertile agreement, and quadratic weighted kappa.

#### eTables

**eTable 1** Baseline predictors used in the primary analysis.

| Variable | Description / Coding | Type |
|---|---|---|
| Age | Continuous, years | Continuous |
| LivingSituation | 3 categories | Categorical |
| Education | 6 categories | Categorical |
| RaceEthnicity | Black / Hispanic | Binary |
| Diabetes | Yes / No | Binary |
| Smoking | Current / Former / Never | Categorical |
| AlcUse | Current / Former / Never | Categorical |
| FamHistHA | Yes / No | Binary |
| HDL_mmolL | mmol/L | Continuous |
| LDL_mmolL | mmol/L | Continuous |
| eGFR_CKD | mL/min/1.73m^2 | Continuous |
| Hemoglobin | g/dL | Continuous |
| SBPMean | mmHg | Continuous |
| DBPMean | mmHg | Continuous |
| BMI | Underweight / Normal / Overweight / Obese | Categorical |
| AbdCirc | Cm | Continuous |
| AvgDominantGrpStr | Dominant hand grip strength | Continuous |
| AvgGaitSpd | Gait speed | Continuous |
| Antihypertensives | Yes / No | Binary |
| LipidLoweringAgents | Yes / No | Binary |
| X3MS_Overall | 3MS score | Continuous |
| CESD_Overall | CES-D score | Continuous |

**eTable 2** Model parameters for the primary Random Survival Forest–based and sensitivity Random Forest–based X-learners.

| Parameter | Primary RSF-based X-learner | Sensitivity RF-based X-learner |
|---|---|---|
| Model type | μ: survival; τ: regression | μ: probability; τ: regression |
| Number of trees | μ: 300; τ: 200 | μ: 300; τ: 200 |
| mtry | μ: 22; τ: 3 | μ: 16; τ: 22 |
| Minimum node size | μ: 3; τ: 2 | μ: 5; τ: 10 |
| Sample fraction | μ: 1.000; τ: 0.632 | μ: 0.800; τ: 0.800 |
| Replacement | μ: no; τ: no | μ: no; τ: yes |
| Split rule | μ: logrank; τ: variance | μ: gini; τ: variance |
| Unordered factors | μ: order; τ: order | μ: partition; τ: ignore |
| Number of predictors | 22 | 22 |

**eTable 3** Seed-based stability of estimated CATEs and predicted benefit tertile assignment. The X-learner was refit using different random seeds while preserving the same analytic cohort, baseline predictors, cross-fitting structure, and model specification. Spearman correlation is used to compare participant-level CATE rankings from each refit with the reference model. Same tertile assignment shows the percentage of participants assigned to the same predicted benefit tertile as in the reference model. Weighted kappa is used to quantify the agreement in tertile assignment beyond chance using quadratic weights.

| Seed | Spearman CATE correlation | Same tertile, % | Weighted κ |
|---|---|---|---|
| 100 | 0.88 | 71.81 | 0.78 |
| 523 | 0.88 | 72.99 | 0.79 |
| 1230 | 0.88 | 72.68 | 0.78 |
| 2129 | 0.89 | 74.49 | 0.81 |
| 2053 | 0.89 | 75.75 | 0.80 |
| 2789 | 0.89 | 75.20 | 0.80 |
| 3055 | 0.88 | 73.39 | 0.79 |
| 4023 | 0.88 | 73.39 | 0.79 |
| 5231 | 0.88 | 74.65 | 0.80 |
| 6481 | 0.89 | 75.75 | 0.81 |

**eTable 4** Discrimination of the arm-specific outcome models at the 5-year horizon.

| Model | AUC |
|---|---|
| Observed-arm prediction | 0.572 |
| Aspirin arm outcome model | 0.632 |
| Placebo arm outcome model | 0.528 |

**eTable 5.** Cox proportional hazards regression results evaluating treatment (trt), CATE (cate_z), and their interaction with the hazard of the outcome.

| Term | HR | 95% CI | SE (log-HR) | Statistic[1] | p-value |
|---|---|---|---|---|---|
| **Cox model coefficients** | | | | | |
| trt | 0.61 | 0.42 to 0.90 | 0.20 | −2.50 | 0.0124 |
| cate_z | 0.78 | 0.62 to 0.98 | 0.12 | −2.16 | 0.0309 |
| trt:cate_z | 1.82 | 1.28 to 2.60 | 0.18 | 3.31 | 0.0009 |
| **Interaction likelihood ratio test** | | | | | |
| trt x cate_z | NA | NA | NA | 10.38 | 0.0013 |

[1] Statistic is Wald z for coefficient rows and likelihood-ratio chi-square for the interaction test row.

**eTable 6** Sensitivity analysis comparing Random Survival Forest and Random Forest–based X-learner results for the US Black and Hispanic cohort.

| Analysis | Scale | Test / Estimate | Primary RSF-based X-learner | Sensitivity RF-based X-learner |
|---|---|---|---|---|
| Continuous CATE interaction | Relative | Interaction p-value (LRT, Cox model) | 0.001 | 0.002 |
| Continuous CATE interaction | Absolute | Interaction p-value (ANOVA, linear model) | 0.011 | 0.006 |
| Grouped CATE interaction (T1– T3) | Relative | Interaction p-value (LRT, Cox model) | 0.011 | 0.025 |
| Grouped CATE interaction (T1– T3) | Absolute | Interaction p-value (ANOVA, linear model) | 0.036 | 0.034 |
| T1 | Relative | HR (95% CI), aspirin vs placebo | 0.36 (0.19 to 0.71) | 0.33 (0.18 to 0.62) |
| T1 | Absolute | ARD at 5 years, percentage points (95% CI) | −11.1 (−22.0 to −0.1) | −13.7 (−24.5 to −2.9) |
| T2 | Relative | HR (95% CI), aspirin vs placebo | 0.48 (0.22 to 1.03) | 0.83 (0.41 to 1.66) |
| T2 | Absolute | ARD at 5 years, percentage points (95% CI) | −4.9 (−13.6 to 3.8) | −1.2 (−10.0 to 7.7) |
| T3 | Relative | HR (95% CI), aspirin vs placebo | 1.26 (0.70 to 2.27) | 1.04 (0.55 to 1.97) |
| T3 | Absolute | ARD at 5 years, percentage points (95% CI) | +3.9 (−5.9 to 13.6) | +1.6 (−8.1 to 11.4) |

#### eFigures

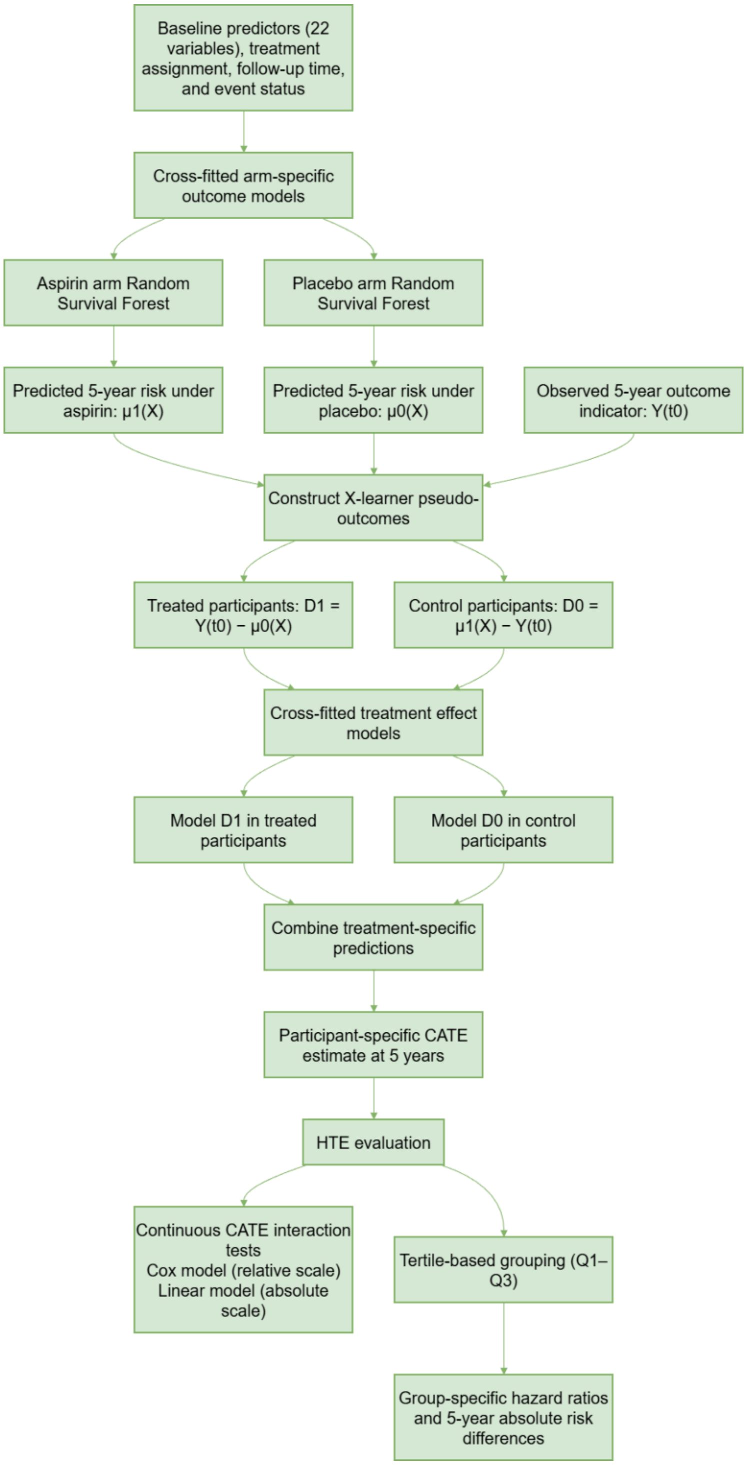


**eFigure 1** Analytic workflow for estimating individualized treatment effects using a Random Survival Forest–based X-learner.

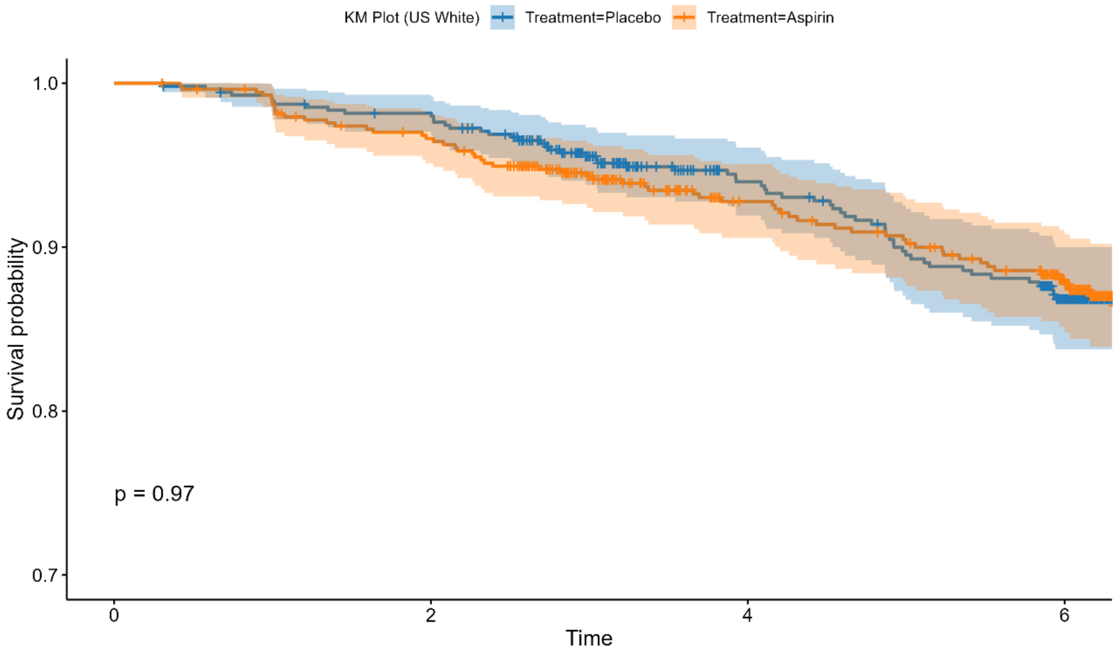


**eFigure 2** Kaplan-Meier survival curves for US non-Hispanic White ASPREE participants comparing aspirin versus placebo.

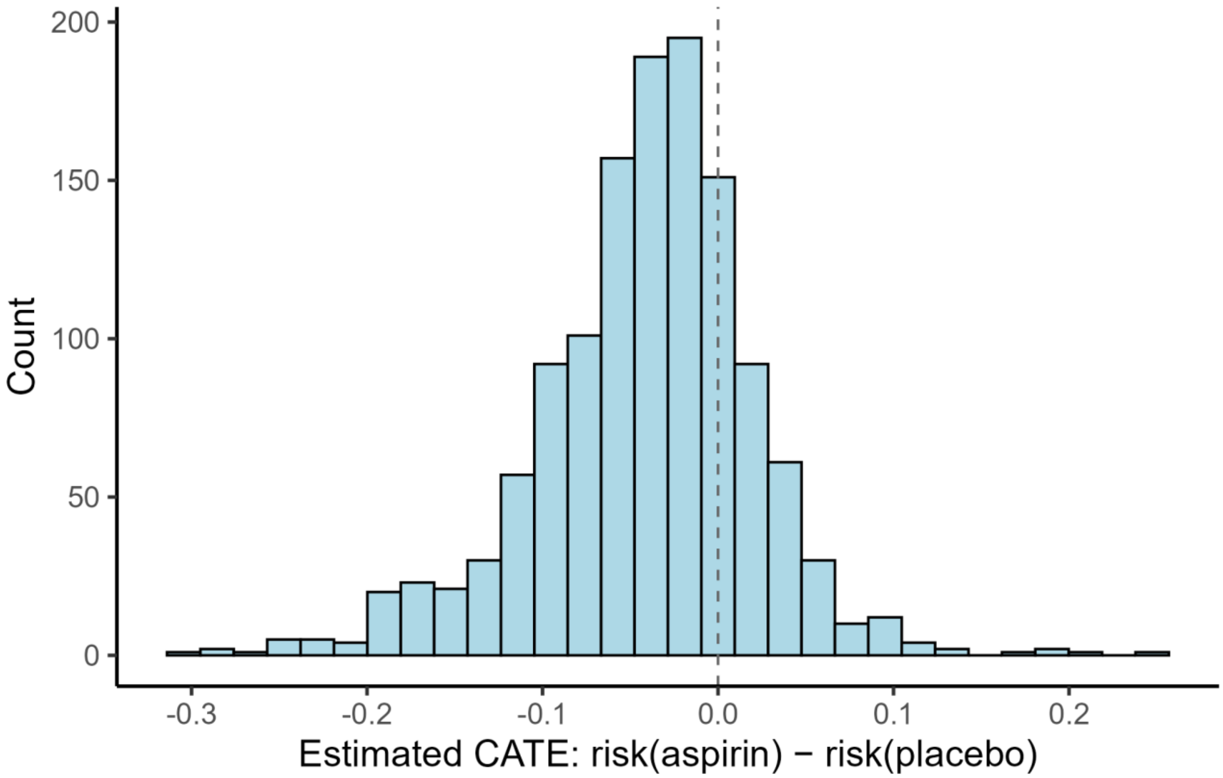


**eFigure 3** Distribution of estimated CATE values from the primary Random Survival Forest–based X-learner among US Black and Hispanic participants.

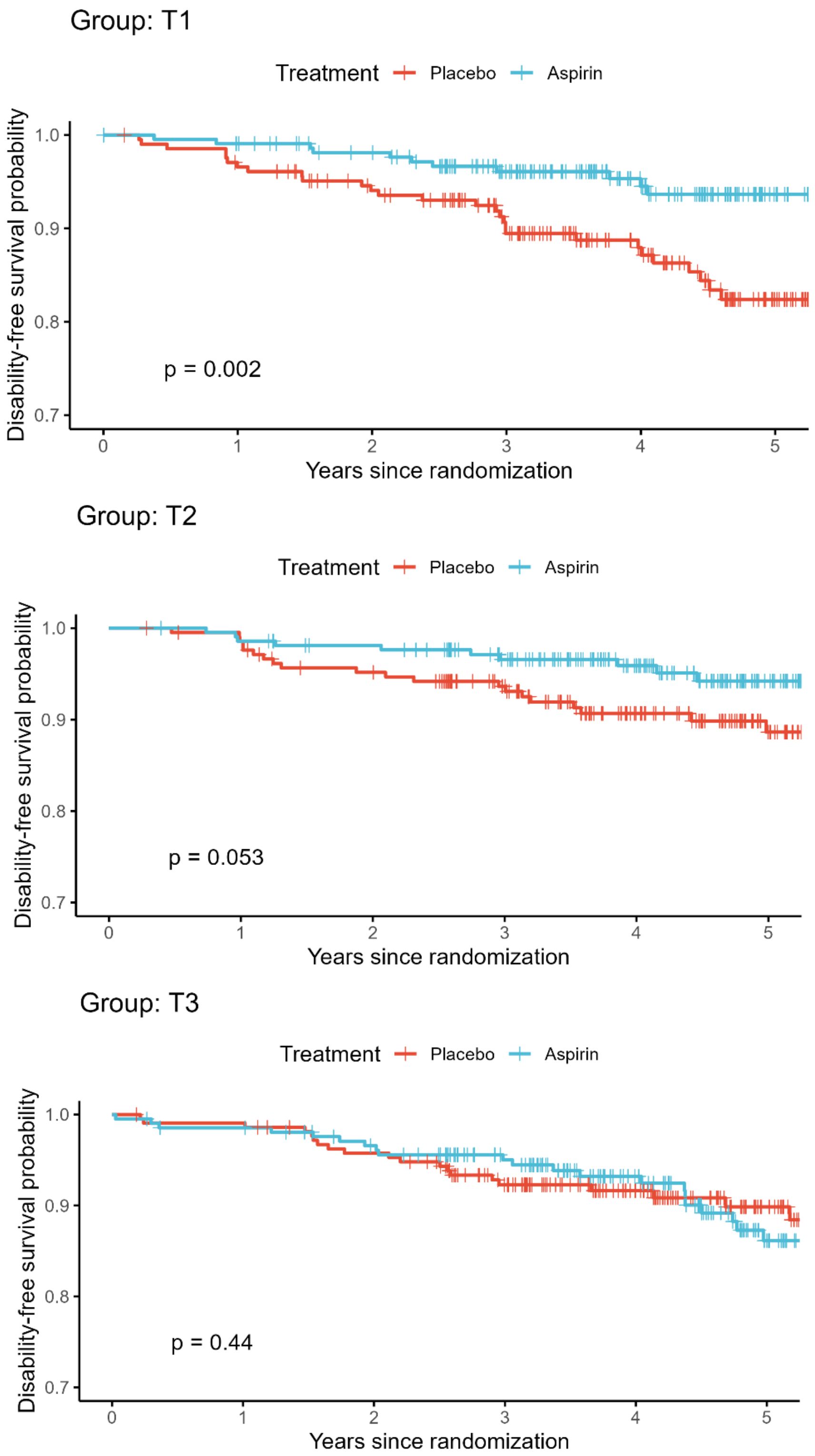


**eFigure 4** Kaplan–Meier curves for disability-free survival stratified by model-derived CATE tertiles and treatment assignment among US Black and Hispanic participants.
